# High SARS-CoV-2 seroprevalence in health care workers but relatively low numbers of deaths in urban Malawi

**DOI:** 10.1101/2020.07.30.20164970

**Authors:** Marah G. Chibwana, Khuzwayo C. Jere, Raphael Kamn’gona, Jonathan Mandolo, Vincent Katunga-Phiri, Dumizulu Tembo, Ndaona Mitole, Samantha Musasa, Simon Sichone, Agness Lakudzala, Lusako Sibale, Prisca Matambo, Innocent Kadwala, Rachel L. Byrne, Alice Mbewe, Marc Y. R. Henrion, Ben Morton, Chimota Phiri, Jane Mallewa, Henry C Mwandumba, Emily R. Adams, Stephen B. Gordon, Kondwani C. Jambo

**Affiliations:** Malawi-Liverpool-Wellcome Trust Clinical Research Programme, Blantyre, Malawi; Centre for Global Vaccine Research, Institute of Infection, Veterinary Ecological Sciences, University of Liverpool, Liverpool, UK; Department of Medicine, College of Medicine, University of Malawi, Blantyre, Malawi; Liverpool School of Tropical Medicine, L3 5QA, Liverpool, United Kingdom; Ministry of Health, Queen Elizabeth Central Hospital, Blantyre, Malawi

**Keywords:** SARS-CoV-2, COVID-19, Malawi, Seroprevalence, IgG

## Abstract

**Background:** In low-income countries, like Malawi, important public health measures including social distancing or a lockdown have been challenging to implement owing to socioeconomic constraints, leading to predictions that the COVID-19 pandemic would progress rapidly. However, due to limited capacity to test for severe acute respiratory syndrome coronavirus 2 (SARS-CoV-2) infection, there are no reliable estimates of the true burden of infection and death. We, therefore, conducted a SARS-CoV-2 serosurvey amongst health care workers (HCWs) in Blantyre city to estimate the cumulative incidence of SARS-CoV-2 infection in urban Malawi.

**Methods:** We recruited 500 otherwise asymptomatic HCWs from Blantyre City (Malawi) from 22^nd^ May 2020 to 19^th^ June 2020 and serum samples were collected from all participants. A commercial ELISA was used to measure SARS-CoV-2 IgG antibodies in serum.

**Results:** A total of 84 participants tested positive for SARS-CoV-2 antibodies. The HCWs with positive SARS-CoV-2 antibody results came from different parts of the city. The adjusted seroprevalence of SARS-CoV-2 antibodies was 12.3% [CI 8.2 - 16.5]. Using age-stratified infection fatality estimates reported from elsewhere, we found that at the observed adjusted seroprevalence, the number of predicted deaths was eight times the number of reported deaths.

**Conclusions:** The high seroprevalence of SARS-CoV-2 antibodies among HCWs and the discrepancy in the predicted versus reported deaths suggests that there was early exposure but slow progression of COVID-19 epidemic in urban Malawi. This highlights the urgent need for development of locally parameterised mathematical models to more accurately predict the trajectory of the epidemic in sub-Saharan Africa for better evidence-based policy decisions and public health response planning.

## Introduction

Coronavirus disease 2019 (COVID-19) has had a dramatic impact worldwide, with high mortality in Asia, Europe and the Americas (1). Africa reported its first COVID-19 case on 14^th^ February 2020 (2). Due to poor socio-economic conditions, high HIV prevalence, an increase in non-communicable diseases and challenged health system infrastructure, it was predicted that the African pandemic would progress rapidly. As of 16^th^ July 2020, however, the number of COVID-19 cases was 665,522 and deaths 14,434 (1, 2), much lower than predicted by mathematical models (3).

In low-income countries, like Malawi, important public health measures like social distancing or a lockdown are difficult to implement owing to socioeconomic constraints. Furthermore, the limited capacity to test for severe acute respiratory syndrome coronavirus 2 (SARS-CoV-2) infection impedes effective public health response planning. Initial testing in Malawi focused on case identification in patients with COVID-19-like symptoms, contacts of index patients and inbound travellers. The first COVID-19 case in Malawi was reported on 2^nd^ April and as of 16^th^ July 2020 there were 2716 cases with only 51 deaths reported (2, 4). Given the sampling strategy, the true burden is certainly much greater than the reported cases, but there are no reliable estimates of the true burden of infection and death. Up to now, health services have reported only small number of cases and have not been overwhelmed as predicted (3).

The unrestricted nature of the COVID-19 epidemic in Malawi provides an opportunity to compare its trajectory in a low-income setting with what has been reported in high income settings. It has been shown that the rate of asymptomatic SARS-CoV-2 infection among health care workers (HCWs) reflects general community transmission rather than in-hospital exposure (5). We, therefore, conducted a SARS-CoV-2 serosurvey amongst HCWs in Blantyre city to estimate the cumulative incidence of SARS-CoV-2 infection in urban Malawi.

## Methods

### Ethical statement

Ethical approval was provided by the College of Medicine Research and Ethics Committee (COMREC, Malawi) (P.05/20/3045) and Liverpool School of Tropical Medicine (LSTM, UK) (20-043). All participants gave informed written consent.

### Study setting and participants

Participant recruitment was done at the Malawi-Liverpool-Wellcome Trust Clinical Research programme (MLW) in Blantyre, Malawi, as part of an ongoing longitudinal study that seeks to investigate markers of SARS-CoV-2 exposure and immunological protection in Malawian adults. The study site is within the compound of the Queen Elizabeth Central Hospital (QECH), the largest tertiary teaching hospital in Malawi. It is easily accessible from most parts of the city. Participants were HCWs from Blantyre City, both clinical and non-clinical. We used a convenience sampling approach, whereby the study was advertised electronically and by word of mouth. The sample size was calculated based on the study primary objective, which was to compare the SARS CoV-2 neutralising antibody titers in recovered COVID-19 patients compared to SARS CoV-2 antibody positive (asymptomatic/mild) individuals. Inclusion criteria for the study included being an HCW resident in Blantyre, aged between 18 and 65 years old, and otherwise symptomatic. The exclusion criterion was withholding consent. Electronic case report forms (eCRFs) were used to collect demographic data including age, gender, place of residence, common mode of transportation, occupation and involvement in COVID-19 work. The samples used for this manuscript are from the baseline recruitment arm of the asymptomatic group in the main study.

### Sample collection, processing and experimental setup

Peripheral blood (10ml), 7ml in Sodium Heparin tubes and 3ml in serum separation tubes (SST) (All BD Biosciences), was collected from all study participants using venesection by the study clinical team at the study site (MLW) between 22^nd^ May 2020 to 19^th^ June 2020. Serum was collected from the SSTs by centrifugation at 500g for 8 mins and stored at -80°C. To measure SARS-CoV-2 antibodies, we used a commercial enzyme linked-immunosorbent assay (ELISA) targeting Spike (S2) and Nucleoprotein (N) from SARS-CoV-2 (Omega diagnostics, UK; ODL 150/10; Lot #103183). The assay was performed as per the manufacturer’s instructions. In brief, participant serum was diluted (1:200) in sample diluent (150mM Tris-buffered saline, pH 7.2 with antimicrobial agent). The diluted samples, diluent alone (negative control), manufacturer’s cut-off control and positive control were added at 100μl per well. The plate was incubated at room temperature for 30 mins. After incubation, the plate was washed three times with wash buffer (100mM Tris-buffered saline with detergent, pH 7.2) using a plate washer (Asys Atlantis, Biochrom Ltd, UK). Human IgG conjugated to horseradish peroxidase was then added to each well at 100μl and incubated for 30 minutes at room temperature. After incubation, the plate was washed four times with wash buffer, and 100μl of TMB (3,3’,5,5’-Tetramethylbenzidine) Substrate (aqueous solution of TMB and hydrogen peroxide) was added. The plate was incubated for 10 minutes at room temperature, before addition of 100μl of Stop Solution (0.25M sulphuric acid). The optical density (OD) of each well was read at 450nm in a microplate reader (BioTek ELx808, UK) within 10 minutes. The assay interpretation was as follows; positive result (OD 0.6), indeterminate result (OD 0.55 to < 0.6) and negative (OD < 0.55) This assay has undergone rigorous independent validation at the Liverpool School of Tropical Medicine (UK) and St George’s University of London (UK) (6).

### Statistical analysis

Graphical presentation was performed using GraphPad Prism 8 (GraphPad Software, USA). To integrate uncertainty arising from test sensitivity and specificity, we used a method published by Reiczigel *et al*, 2010 (7). The geospatial data was plotted in R (v4.0.0) using ggmap (v3.0.0) (8) and ggplot2 (v3.3.1) (9).

## Results

### Demographics of study participants

We recruited 500 asymptomatic HCWs with a median age of 31 (range 20-64 years). The average household size for the participants was 4 [confidence interval (CI) 3–5]. Of the 500, 331 were clinical HCWs and 169 non-clinical HCWs (Table 1). The clinical HCWs included nurses, medical doctors and clinical officers, while the non-clinical HCWs included clerical/administration, field workers and laboratory scientists. The primary workstation for the HCWs included primary healthcare facilities (35/500), secondary healthcare facilities (291/500), and clinical research facilities (174/500). The majority of the participants were nurses (57%), 41% of all participants were involved in clinical work related to COVID-19 and 73% of the total participants used public transport or walking as their main means of transport. The main characteristics of the participants are summarised in Table 1.

**Table 1.** Study participant demographics

| Characteristics | Number of participants | Proportion |
| --- | --- | --- |
| **Gender** |  |  |
| Male | 236 | 47% |
| Female | 264 | 53% |
| **Age** |  |  |
| 20-29 | 209 | 42% |
| 30-39 | 170 | 34% |
| 40-49 | 86 | 17% |
| 50-59 | 28 | 6% |
| 60+ | 7 | 1 |
| **Occupation** |  |  |
| Clinical HCW | 331 | 66% |
| Non-Clinical HCW | 169 | 34% |
| **COVID-19 work** |  |  |
| Clinical | 205 | 41% |
| Non-Clinical | 43 | 9% |
| None | 252 | 50% |
| **Transport** |  |  |
| Public + Walking | 363 | 73% |
| Private | 137 | 27% |

**COVID-19, Coronavirus Diseases of 2019; HCW, Health Care Workers**

### Seroprevalence of SARS CoV-2 antibodies and geospatial location of the antibody positive individuals

84 participants tested positive for SARS-CoV-2 antibodies (Figure 1). After adjusting for test sensitivity and specificity (6, 7), the overall seroprevalence of SARS-CoV-2 antibodies was 12.3% [CI 8.2 - 16.5]. This suggests that local transmission was high and that SARS-CoV-2 may have been circulating for some time in Blantyre City.

**Figure 1.**
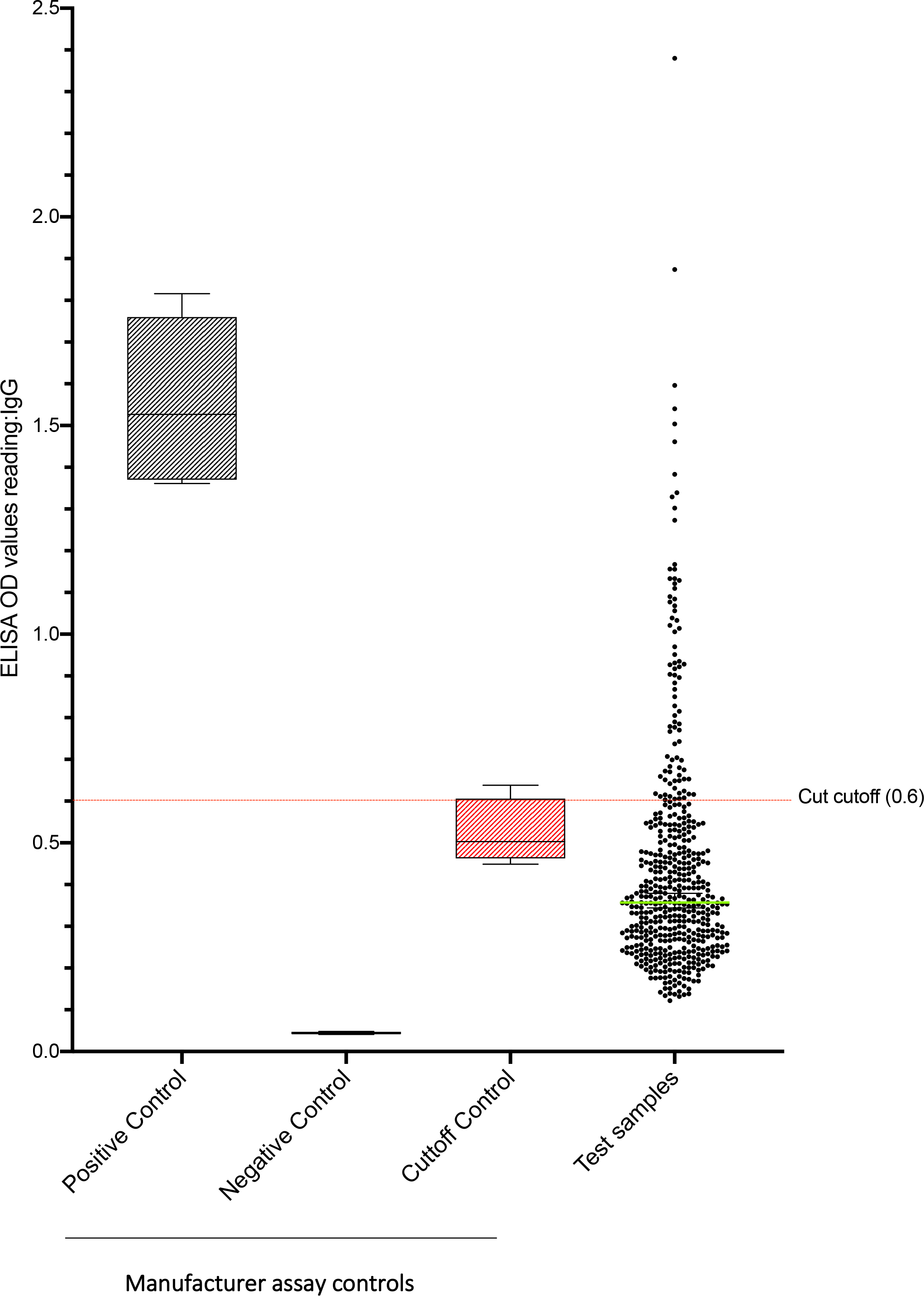
SARS-CoV-2 serological results from asymptomatic health care workers. We used a commercial ELISA to measure SARS-CoV-2 antibodies against Spike (S2) and Nucleoprotein (N). OD, optical density.

To estimate the potential geographical spread of SARS-CoV-2, we plotted the geographical coordinates of place of residence for the individuals with a positive antibody result on the map of Blantyre City. We found that the HCWs with a positive SARS-CoV-2 antibody result came from different parts of the city (Figure 2). This suggests that SARS-CoV-2 local transmission was likely widespread across the city.

**Figure 2.**
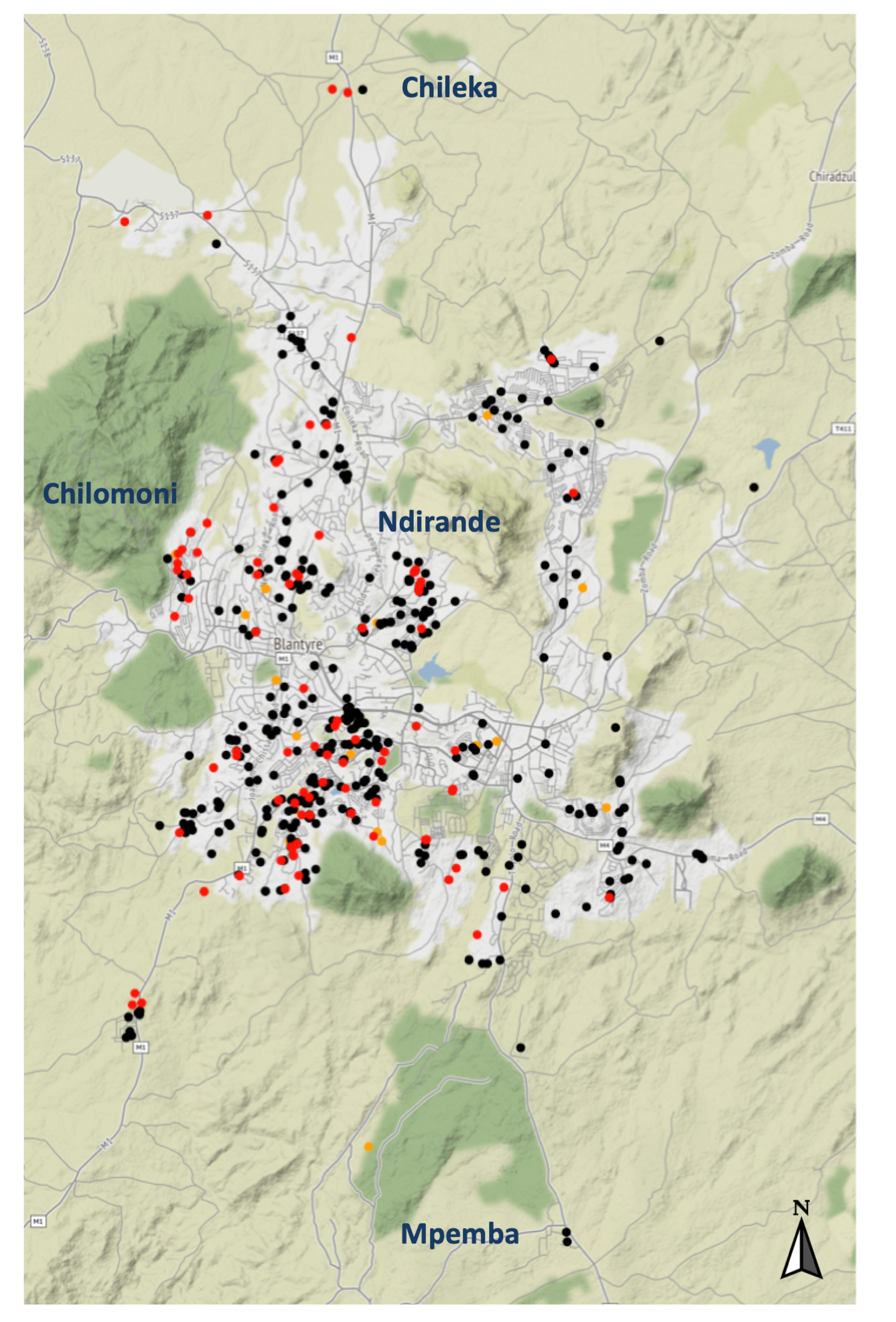
Map of Blantyre showing geospatial distribution of seropositive results. We collected geocoordinate data for the place of residence of all study participants at recruitment. The geocoordinates were combined with the ELISA assay results and plotted on the map of Blantyre using R. Black dot, seronegative; Orange dot, indeterminate; Red dot, seropositive.

### Crude projections of mortality based on seroprevalence estimates

Using estimates of infection fatality rates from Verity *et al*. (10) and the Malawi population census (11), we estimated the number of deaths that could have occurred at the observed seroprevalence of SARS-CoV-2 antibodies (Table 2). We adjusted the population estimates by inflating them to take into account population annual population growth rate of 2% from 2018 to 2020 (11). We assumed that there was a uniform risk of infection at all age groups and that the seroprevalence was similar to the general population.

**Table 2.**
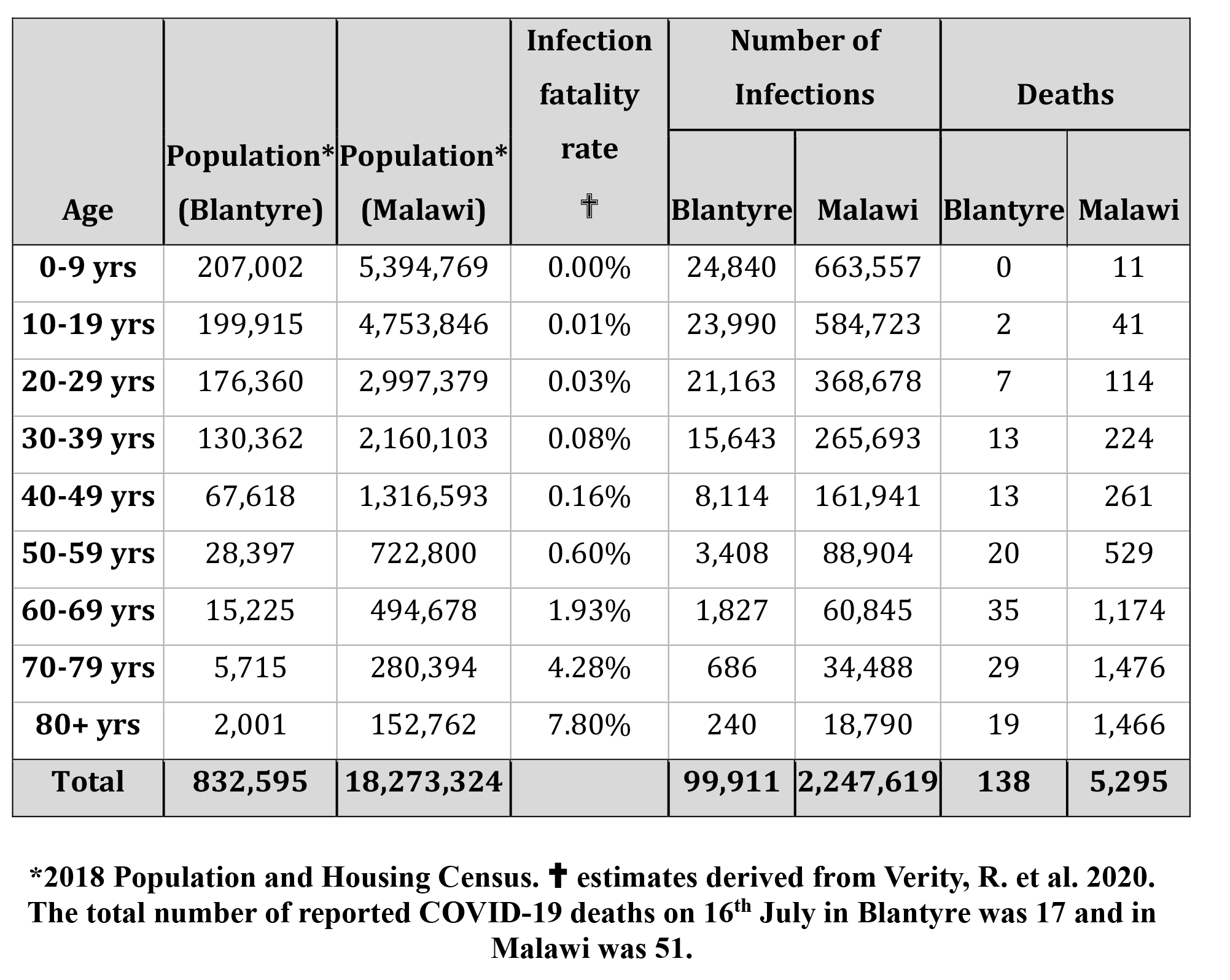
Crude estimates of predicted mortality at the observed seroprevalence

The crude estimates suggest that there should have been at least 138 deaths by 19^th^ June 2020. However, four weeks following the serosurvey, only 17 COVID-confirmed deaths in Blantyre have been reported by the Public Health Institute of Malawi (4), which is approximately eight times below the predicted deaths. When the seroprevalence is extrapolated to the entire Malawi, it predicts approximately 5,295 COVID-19 deaths, but only 51 deaths have been reported as of 16^th^ July 2020. These crude estimates highlight a discrepancy between the predicted deaths using infection fatality rates from elsewhere and the actual number of reported COVID-19 deaths in Malawi.

## Conclusions

To our knowledge, this seroprevalence study is the first to report estimates of SARS-CoV-2 exposure among HCWs in an African urban low-income setting. It provides insights into the potentially unique trajectory of the COVID-19 epidemic in sub-Saharan Africa (SSA), using data from urban Malawi. We observe a high seroprevalence of SARS-CoV-2 antibodies amongst HCWs. It has been reported elsewhere that HCWs accounted for a high proportion of cases early in the SARS-CoV-2 outbreak when transmission was increasing sharply and personal protective equipment (PPE) provision was patchy (12-14). Our data could suggest that Malawi is relatively early in the epidemic and that COVID-19 cases are likely to continue to rise sharply in the coming weeks, but the serology also suggests that large numbers of cases must be either asymptomatic or only show mild disease.

The discrepancy between the predicted compared to reported mortality at the observed seroprevalence estimate may also suggest that there are large numbers of underreported or misclassified deaths in Malawi. However, even in countries like South Africa with relatively abundant testing capacity and strong health systems, there is relatively low mortality with a case fatality ratio of 1.5 (15). This may imply that the impact of SARS-CoV-2 in Africa is potentially much less severe or is following a different trajectory than that experienced in China, Americas and Europe, where case fatality ratios were commonly above five (1). This warrants further investigation.

However, the reasons behind the discrepancy in the COVID-19 pandemic trajectory between SSA and elsewhere might include population demography, climate and prior cross-reactive immunity (16). In Malawi, for example, the population is younger (median age 17 years old) (11), and the elderly who mostly experience worse outcomes in other settings (10), are 5.1% of the population (11), largely residing in rural areas. If the prevalence of SARS-CoV-2 is very low in rural areas, this may explain the low number of deaths, and would strengthen the call to shield the elderly (17).

This study has some limitations. First, selection bias is likely due to the convenience sampling approach; however, targeting HCW for regular serosurveys could help predict local transmission outbreaks. Second, this serosurvey focused on an urban population where Malawi has reported the highest concentration of COVID-19 cases. The seroprevalence in the rural population remains unknown, but if high, may prompt other explanations for the African/Malawi situation. Third, current SARS-CoV-2 ELISAs are still undergoing rigorous validation and verification in the African settings, hence seroprevalence estimates could change with new information on the accuracy of the test kits.

In conclusion, our findings indicate a major discrepancy between predicted COVID-19 mortality at the observed SARS-CoV-2 seroprevalence in HCWs with reported COVID-19 deaths in urban Malawi. The high seroprevalence estimate implies earlier exposure of SARS-CoV-2 than that reported but with slow progression of the COVID-19 epidemic. Development of locally parameterised mathematical models should be prioritised to more accurately predict the trajectory of the epidemic in SSA. This will allow better evidence-based policy decision-making and public health response planning.

## Data Availability

All data is available on Figshare: High SARS-CoV-2 seroprevalence in Health Care Workers but relatively low numbers of deaths in urban Malawi.
This project contains the following underlying data:
- PROTECT_FigShare Part 1 31072020.csv
- PROTECT_screening_enrolment_data_dictionary Figshare 31072020.pdf
Data are available under the terms of the Creative Commons Attribution 4.0 International license (CC-BY 4.0).

https://doi.org/10.6084/m9.figshare.12745214.v2

## Data availability

Figshare: High SARS-CoV-2 seroprevalence in Health Care Workers but relatively low numbers of deaths in urban Malawi. https://doi.org/10.6084/m9.figshare.12745214.v2

This project contains the following underlying data:

‐ PROTECT_FigShare Part 1 31072020.csv
‐ PROTECT_screening_enrolment_data_dictionary Figshare 31072020.pdf

Data are available under the terms of the <u>Creative Commons Attribution 4.0 International license</u> (CC-BY 4.0).

## Acknowledgements

We acknowledge the PROTECT Study Team at the Malawi-Liverpool-Wellcome Trust Clinical Research Programme, namely Alice Kalilani, Gift Sagawa, Martha Moyo, Orpha Kumwenda, Sharon Nthala, Chisomo Jassi, Neema Nyakuleha and Tayamika Banda. We thank Dr Joe Fitchett at Mologic (UK) for providing the SARS-CoV-2 ELISA kits and Dr Peter McPherson (MLW/LSTM) for the epidemiological support.

## Disclaimer

The funders had no role in the study design, data collection and interpretation, or the decision to submit the work for publication. Mologic (UK) was provided the opportunity to review a preliminary version of this manuscript for factual accuracy but the authors are solely responsible for final content and interpretation. The authors received no financial support or other form of compensation related to the development of the manuscript. The findings and conclusions in this report are those of the authors.

